# POPULATION ANALYSIS OF MORTALITY RISK: PREDICTIVE MODELS USING MOTION SENSORS FOR 100,000 PARTICIPANTS IN THE UK BIOBANK NATIONAL COHORT

**DOI:** 10.1101/2022.04.20.22274067

**Authors:** Haowen Zhou, Ruoqing Zhu, Anita Ung, Bruce Schatz

**Affiliations:** Department of Statistics, University of Illinois at Urbana-Champaign, Urbana Illinois 61801 USA; College of Medicine, University of Illinois at Urbana-Champaign, Urbana Illinois 61801 USA; Carl R. Woese Institute for Genomic Biology, University of Illinois at Urbana-Champaign, Urbana Illinois 61801 USA

## Abstract

We have developed novel technology for health monitoring, which inputs motion sensors to predictive models of health status. We have validated these models in clinical experiments with carried smartphones, using only their embedded accelerometers. Using smartphones as passive monitors for population measurement is critically important for health equity, since they are ubiquitous in high-income countries already and will become ubiquitous in low-income countries in the near future. Our study simulates smartphones by using accelerometers as sensor input.

We analyzed 100,000 participants in UK Biobank who wore activity monitors with motion sensors for 1 week. This national cohort is demographically representative of the UK population, and this dataset represents the largest such available sensor record. We performed population analysis using walking intensity, with participants whose motion during normal activities included daily living equivalent of timed walk tests. We extract continuous features from sensor data, for input to survival analysis for predictive models of mortality risk.

Simulating population monitoring, we validated predictive models using only sensors and demographics. This resulted in C-index of 0.76 for 1-year risk decreasing to 0.73 for 5-year. A minimum set of sensor features achieves similar C-index with 0.72 for 5-year risk, which is similar accuracy to previous studies using methods not achievable with phone sensors. The minimum model uses average acceleration, which has predictive value independent of demographics of age and sex, as does the physical measure of gait speed. Our digital health methods achieve the same accuracy as activity monitors measuring total activity, despite using only walking sessions as sensor input, orders of magnitude less than existing methods.

**AUTHOR SUMMARY:** Supporting healthcare infrastructure requires screening national populations with passive monitors. That is, looking for health problems without intruding into daily living. Digital health offers potential solutions if sensor devices of adequate accuracy for predictive models are already widely deployed. The only such current devices are cheap phones, smartphone devices with embedded sensors. This limits the measures to motion sensors when the phones are carried during normal activities. So measuring walking intensity is possible, but total activity is not.

Our study simulates smartphone sensors to predict mortality risk in the largest national cohort with sensor records, the demographically representative UK Biobank. Death is the most definite outcome, accurate death records are available for 100,000 participants who wore sensor devices some five years ago. We analyzed this dataset to extract walking sessions during daily living, then used these to predict mortality risk. The accuracy achieved was similar to activity monitors measuring total activity and even to physical measures such as gait speed during observed walks. Our scalable methods offer a potential pathway towards national screening for health status.

## INTRODUCTION

The association of physical activity with mortality risk is well known. National cohort studies using self-reports have shown intensity is correlated with survival, so persons with lower mortality have more moderate-to-vigorous activity and less sedentary activity [1]. These studies focus upon the volume of activity at certain level of intensity. These have been replicated with large meta-analysis studies using objective physical activity, where wearable sensors record total activity and statistical models predict mortality risk from accelerometer measures [2]. Cohort meta-analysis also shows sensor features improve model performance beyond traditional risk factors [3], e.g. smoking and alcohol, while independent of demographics, e.g. age and sex.

Besides quantity of intensity, there are effects of quality of intensity. Physical measurements focus upon walking as moderate activity, in-between vigorous and sedentary. Large meta-analysis studies show gait speed is correlated with mortality risk [4], with timed walking over short distances such as 6 seconds for 4 meters. National cohort studies using self-reports show walking pace is the unique feature beyond traditional risk factors in improving mortality risk for cardiovascular disease [5]. The 6 minute walk test [6], where persons walk steadily in hospital corridor, is a standard evaluation for cardiopulmonary disease. This test has been shown in large meta-analysis studies to be strong independent predictor of mortality from heart failure [7].

We analyze the largest national cohort, the UK Biobank [8], where 103,683 participants wore accelerometer devices for 1 week as wrist sensors [9]. Following our previous analysis of a national cohort for physical activity in the US Women’s Health Initiative [10], we used raw sensor data during labelled walking sessions to identify characteristic motions for predictive models. This is the first population analysis of walking intensity with mobile sensors, and uses only inputs that could be accurately gathered using only personal smartphones.

As surmised from this introduction, there are four primary methods for measuring physical activity. It will be shown later that all these methods achieve roughly the same accuracy for predictive models of mortality risk. Two methods are active, requiring persons explicitly do some activity, such as answering a questionnaire concerning their health status (self-report) or walking fixed distance under observation (gait-speed). These have proven feasible within cohort studies, but are problematic for population health, due to logistic difficulty of getting large numbers of people to perform the required tasks on a routine basis. Two methods are passive, requiring persons to wear measurement devices, such as activity monitors, to measure total activity during the day or specific measures like walking pace over limited periods. These sensor-based methods have the major advantage that they can measure physical activity in daily living, without requiring persons to change their normal activity other than wearing the devices.

However, such digital health approaches have had limited success due to problems with health equity. Results with activity monitors are largely from representative samples, such as national cohorts as in this study, rather than from actual large populations. For population measurement in health systems to be routinely available, the measurement devices must be widely deployed [11], which requires mobile phones at present day. In the United States for example, the Pew Research Center estimates 97% of the population has cell phones with 83% having smart phones containing motion sensors [12]. But only 21% of the population has wearable sensors within smart watches or fitness devices [13]. So scalable methods for predictive models using mobile phones would have great impact if limitations can be overcome. Mobile phones are often carried while walking, so could passively capture walking sessions with motion sensors, but rarely carried all day, so not effective to measure total amount of physical activity unlike wearables.

The smartphone penetration rate in the United Kingdom, where our dataset was gathered, has increased every year over the past decade, reaching an overall ownership of 92% in 2021. For older adults in the statistical survey [14], less than half of all respondents over the age of 55 owned such a device in 2016, but this total rose to 83% in 2021. So adequate devices will soon be everywhere, when the cheapest flip phones have motion sensors. Cheap phones are already widespread worldwide, even in the poorest countries [15]. The global smartphone penetration rate is estimated to have reached over 78 percent in 2020. This is based on 6.4 billion smartphone subscriptions in a global population of 7.8 billion. The global smartphone penetration rate in the general population has great regional variation. In North America and Europe, the smartphone adoption rate stands at roughly 82 and 78 percent, respectively. Whereas in Sub-Saharan Africa, the same rate only stands at 48 percent as of late, showing a roughly 30 percent difference in adoption rates between the highest and lowest ranked regions. But note that even in low-income regions, half the population already has smart phones with motion sensors.

Since mortality is the definite extreme case of health status, cheap phones could have major impact in addressing health equity if proper models can be developed utilizing only their sensors when they are carried. Our study uses the sensor dataset from the largest current national cohort, the UK Biobank. Although this data was gathered from activity monitors, our sensor models use only the inputs feasible to gather with cheap phones. This is possible because of our extensive clinical experiments with cheap phones, developing highly accurate predictive models for health status with cardiopulmonary patients [16]. The scale of 100K participants provides significant generality to the model results, given that the cohort demographics for all 500K participants in UK Biobank matches the demographics of the national population [17], and their sensor cohort has similarly representative demographics.

## RESULTS

We show short bursts of steady walking suffice for predictive models of mortality risk, evaluated using raw sensor data for 100,000 participants in UK Biobank. Our Results evaluate the model accuracy for mortality risk using walking intensity, defined as 12 walking windows of 30 seconds each during a consecutive session, representing daily living versions of walk tests. Our accuracy is comparable to previous models using daily profiles of activity volume. Our methods are logistically easier, with 6 minutes per day (12 windows) rather than 600 minutes (10 hours) per day of sensor records. Although the analyzed dataset uses wrist sensors, our previous work showed cheap smart phones have good enough accelerometer sensors to be accurately utilized for similar analysis of walking sessions [18]. Our clinical studies have shown predictive models using only walking intensity can accurately compute pulmonary function for cardiopulmonary patients [16]. Thus our analysis with wearable sensors for predicting mortality is directly applicable for clinical practice with personal smartphones, already ubiquitous in the UK and the US populations, and widespread in global populations.

### Max (maximum) Models

To model mortality, we consider maximum follow-up time of 1/2/3/4/5 years. This means when the maximum follow-up time is 1 year, any event after 1 year after sensor records is ignored. So we can evaluate model accuracy in early risk years, as well as standard 5-year mortality. The UK Death Registry is used to determine which participants had died by that time.

As detailed in the Methods section, we choose 20 traditional predictors, from self reports and laboratory tests. These 20 questions are listed in Table 1 as the Categorical Features. The full encoding from UK Biobank data is given in Supplementary Table S1. We also choose 76 derived predictors from motion (accelerometer) sensors. These 76 sensor features are listed in Table 2 as the Continuous Features. The full encoding from UK Biobank software [19] is given in Supplementary Table S2. We fit a penalized Cox proportional hazard model with all these features, and denote this the *Max Model*, which evaluates accuracy with maximum functionality. Figure 1 gives the computation flowchart for predictive models.

**Figure 1.**
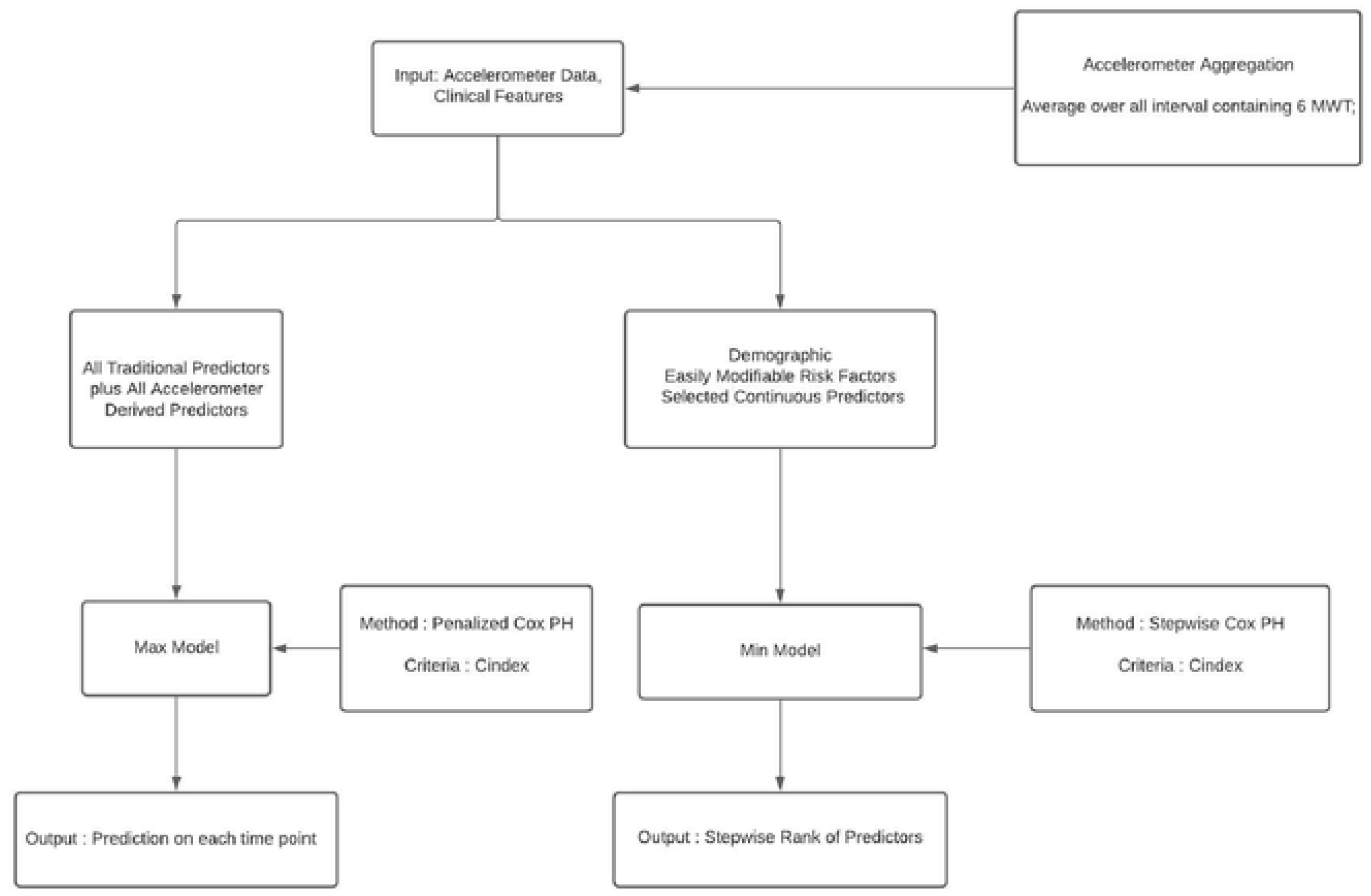
Model Flowchart. Which Inputs used for Which Models.

**Table 1.**
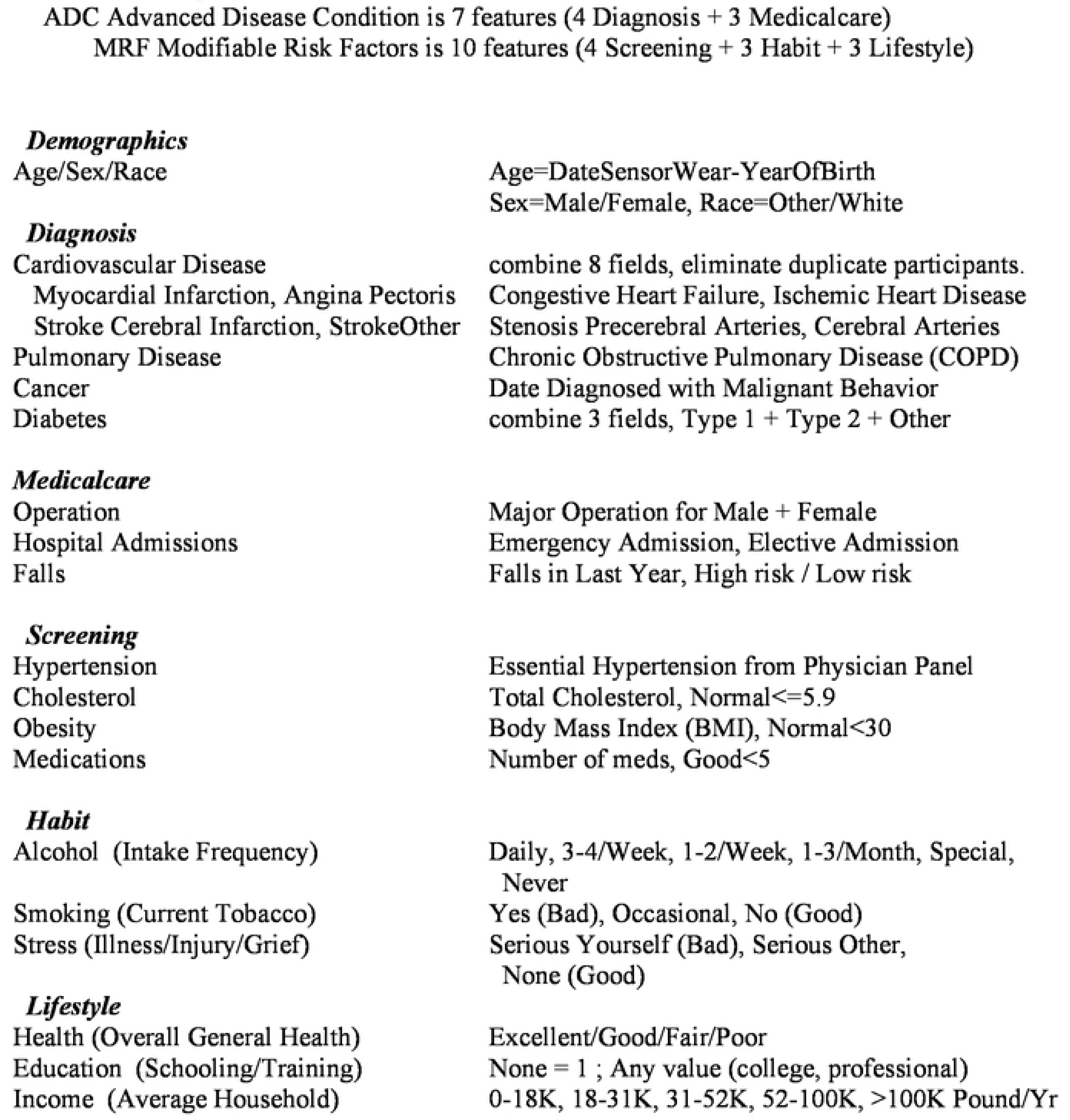
Categorical Features from UK Biobank dataset fields.

**Table 2.**
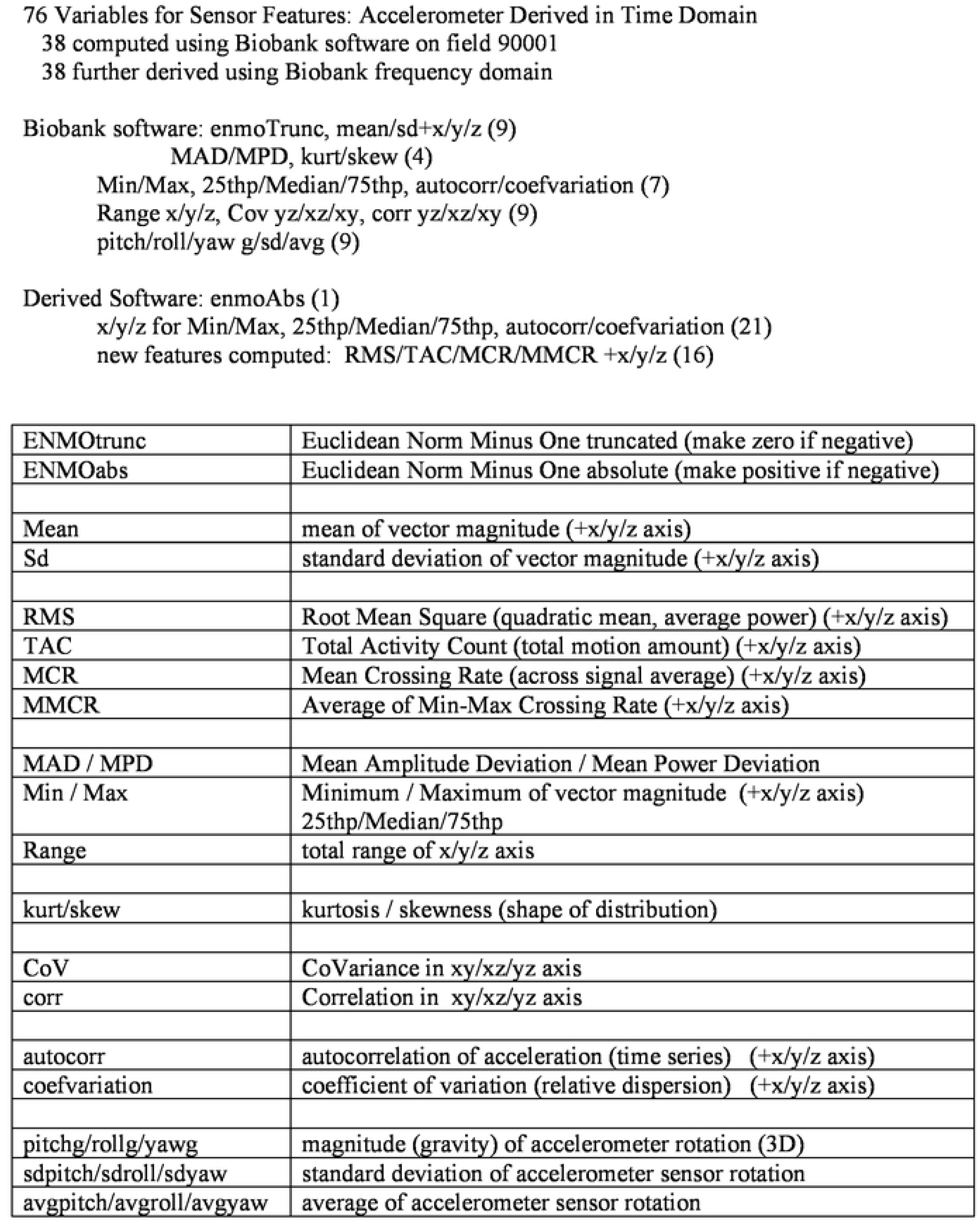
Continuous Features from Participant Sensor Records.

We computed Max Models for different groups of categorical features. All the Models included the 3 demographic features (age/sex/race). The continuous variables are all 76 sensor features, with only steady walking as model input. Figure 2 shows that 100,655 participants met the inclusion criteria of 1 walking session of 6 steady minutes during the 1 week. The Max Model plots are shown in Figure 3. The plot showing continuous variables alone is given in Figure 3a, where continuous is a distinct improvement on demographics. The C-index is 0.76 at 1-year risk, falling to 0.73 at 5-year risk. The modifiable risk factors are similar at 1-year, where the sensors are more recent, but slightly better at 5-year risk. The advanced disease are significantly better even more at 1-year, but converge to the same as risk factor at 5-year risk.

**Figure 2.**
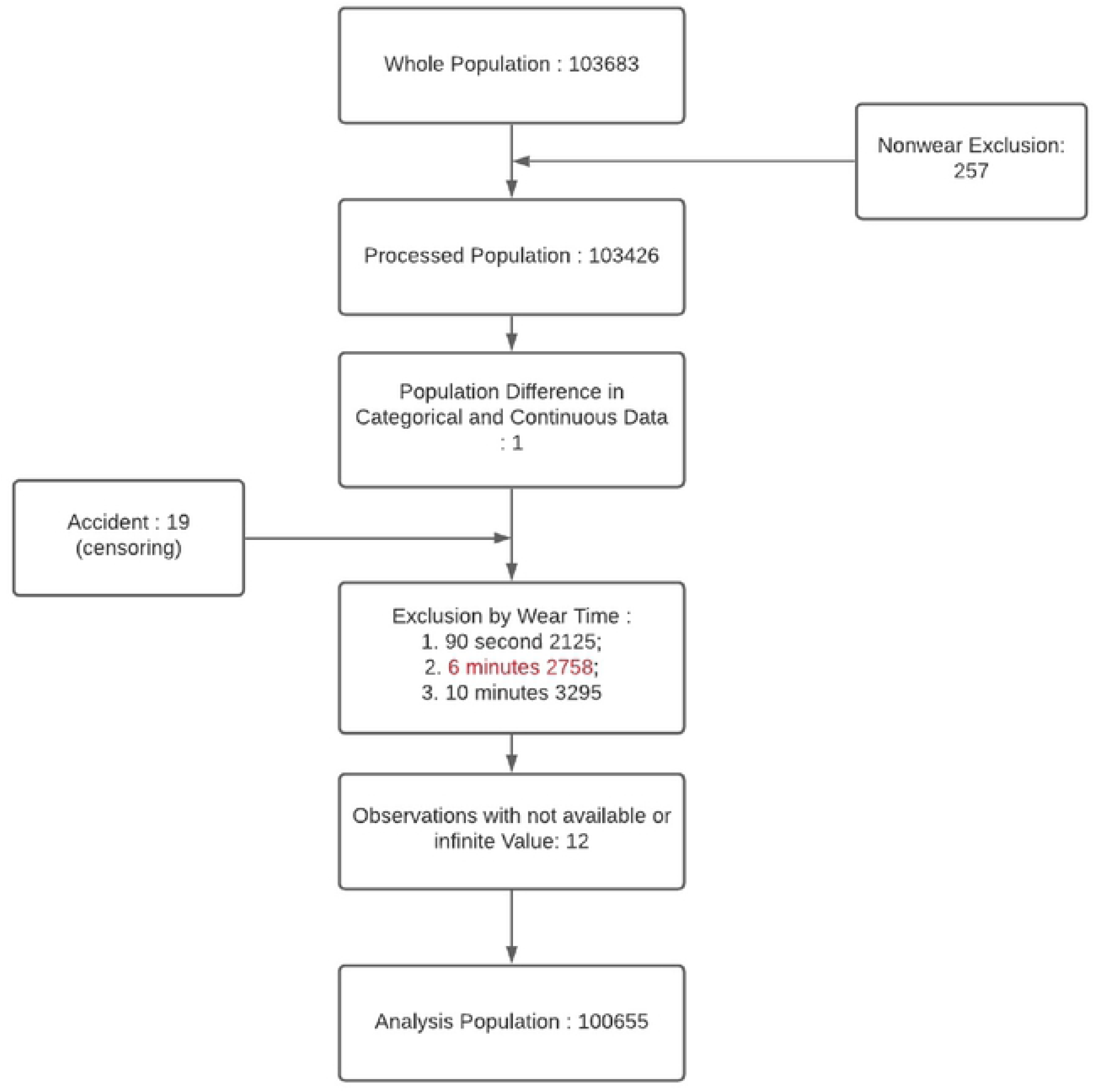
Participant Flowchart. Inclusion/Exclusion for Mortality Prediction.

**Figure 3.**
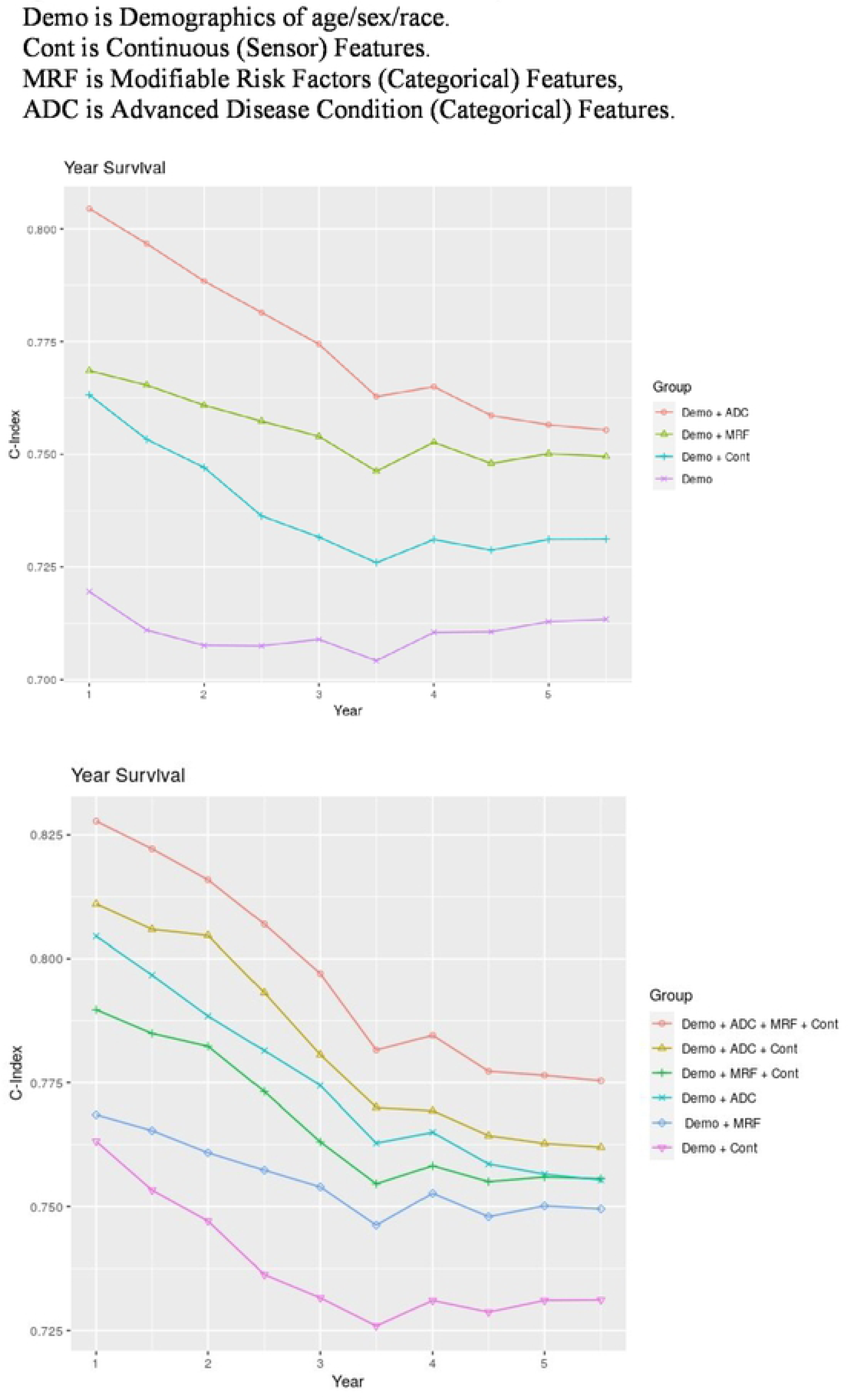
Max Model plots with Demographics and Features curves.

The continuous variables always improve the accuracy of any set of features, as plots show in Figure 3b. Continuous features with demographics only at the bottom is 0.76 at 1-year and 0.73 at 5-year. Whereas continuous with all categorical features at the top is 0.83 at 1-year and 0.78 at 5-year. In-between, continuous slightly improves the curve for risk factors and the curve for advanced disease. The C-index evaluation numbers of all the Max Models are given in Table 3.

**Table 3.**
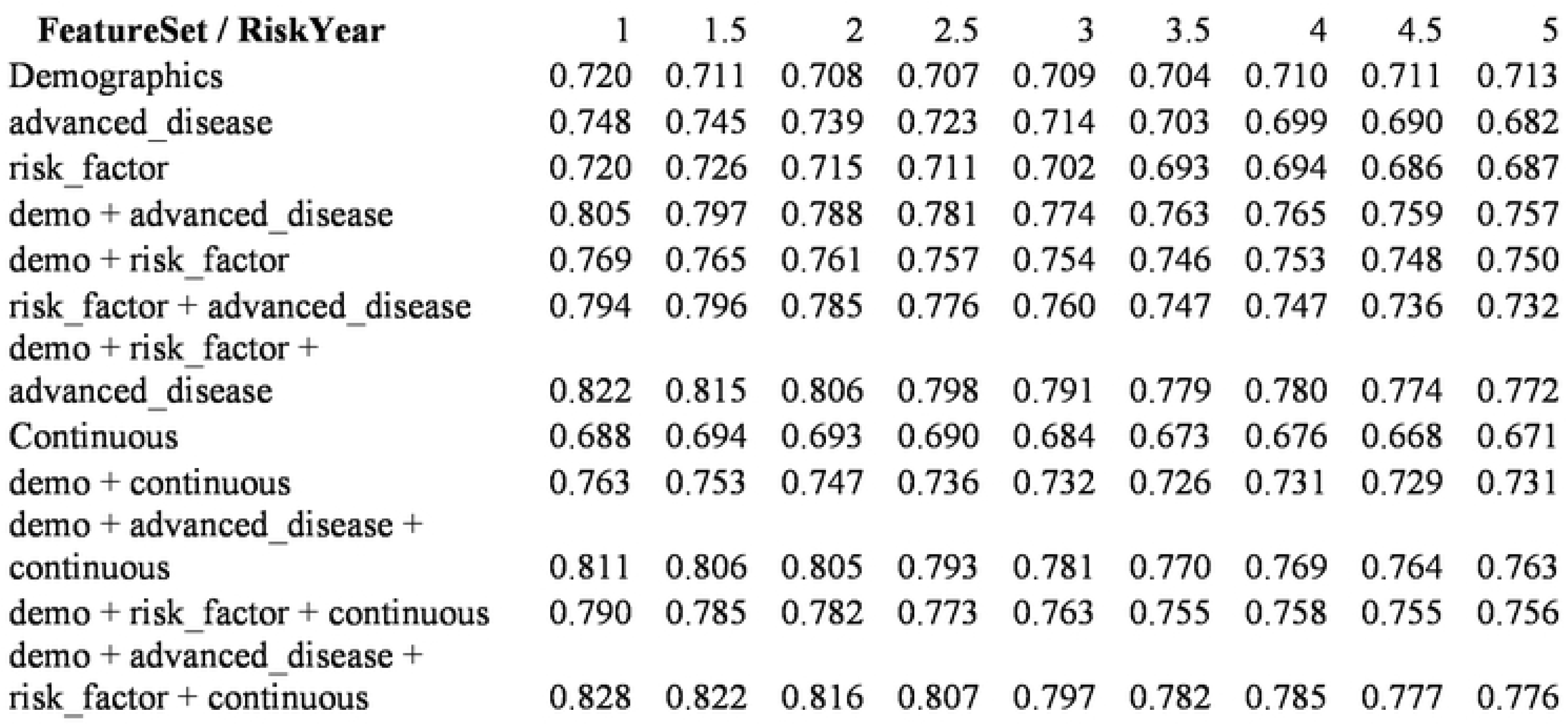
Max Model results with C-index, Feature Sets versus Risk Years.

| **FeatureSet / RiskYear** | 1 | 1.5 | 2 | 2.5 | 3 | 3.5 | 4 | 4.5 | 5 |
| --- | --- | --- | --- | --- | --- | --- | --- | --- | --- |
| Demographics | 0.720 | 0.711 | 0.708 | 0.707 | 0.709 | 0.704 | 0.710 | 0.711 | 0.713 |
| advanced_disease | 0.748 | 0.745 | 0.739 | 0.723 | 0.714 | 0.703 | 0.699 | 0.690 | 0.682 |
| risk_factor | 0.720 | 0.726 | 0.715 | 0.711 | 0.702 | 0.693 | 0.694 | 0.686 | 0.687 |
| demo + advanced_disease | 0.805 | 0.797 | 0.788 | 0.781 | 0.774 | 0.763 | 0.765 | 0.759 | 0.757 |
| demo + risk_factor | 0.769 | 0.765 | 0.761 | 0.757 | 0.754 | 0.746 | 0.753 | 0.748 | 0.750 |
| risk_factor + advanced_disease | 0.794 | 0.796 | 0.785 | 0.776 | 0.760 | 0.747 | 0.747 | 0.736 | 0.732 |
| demo + risk_factor + advanced_disease | 0.822 | 0.815 | 0.806 | 0.798 | 0.791 | 0.779 | 0.780 | 0.774 | 0.772 |
| Continuous | 0.688 | 0.694 | 0.693 | 0.690 | 0.684 | 0.673 | 0.676 | 0.668 | 0.671 |
| demo + continuous | 0.763 | 0.753 | 0.747 | 0.736 | 0.732 | 0.726 | 0.731 | 0.729 | 0.731 |
| demo + advanced_disease + continuous | 0.811 | 0.806 | 0.805 | 0.793 | 0.781 | 0.770 | 0.769 | 0.764 | 0.763 |
| demo + risk_factor + continuous | 0.790 | 0.785 | 0.782 | 0.773 | 0.763 | 0.755 | 0.758 | 0.755 | 0.756 |
| demo + advanced_disease + risk_factor + continuous | 0.828 | 0.822 | 0.816 | 0.807 | 0.797 | 0.782 | 0.785 | 0.777 | 0.776 |

Continuous variables have many similarities. Sensor features extracted from signal processing on raw accelerometry compute the same input with similar outcomes on predictive models. Each feature does provide additional accuracy, as shown in Supplementary Table S3, which gives marginal performance of each feature by itself. The top features after Age are ENMOtrunc and MPD, along with their variant computations ENMOabs and MAD. These are features measuring the average acceleration of sensor signal via Euclidean Norm or Mean Deviation. ENMO is Euclidean Norm Minus One, after adjusting acceleration for effects of gravity [20].

Lasso models can be utilized to select fewer features, by focusing on those most predictive [21]. The additional discrimination provided by each feature flattens out quickly, so 4 features provide the same model accuracy as all 76 features, for cross-validated C-index. This is shown in Figure 4, which shows cumulative effort of multiple features flattening after 4 features.

**Figure 4.**
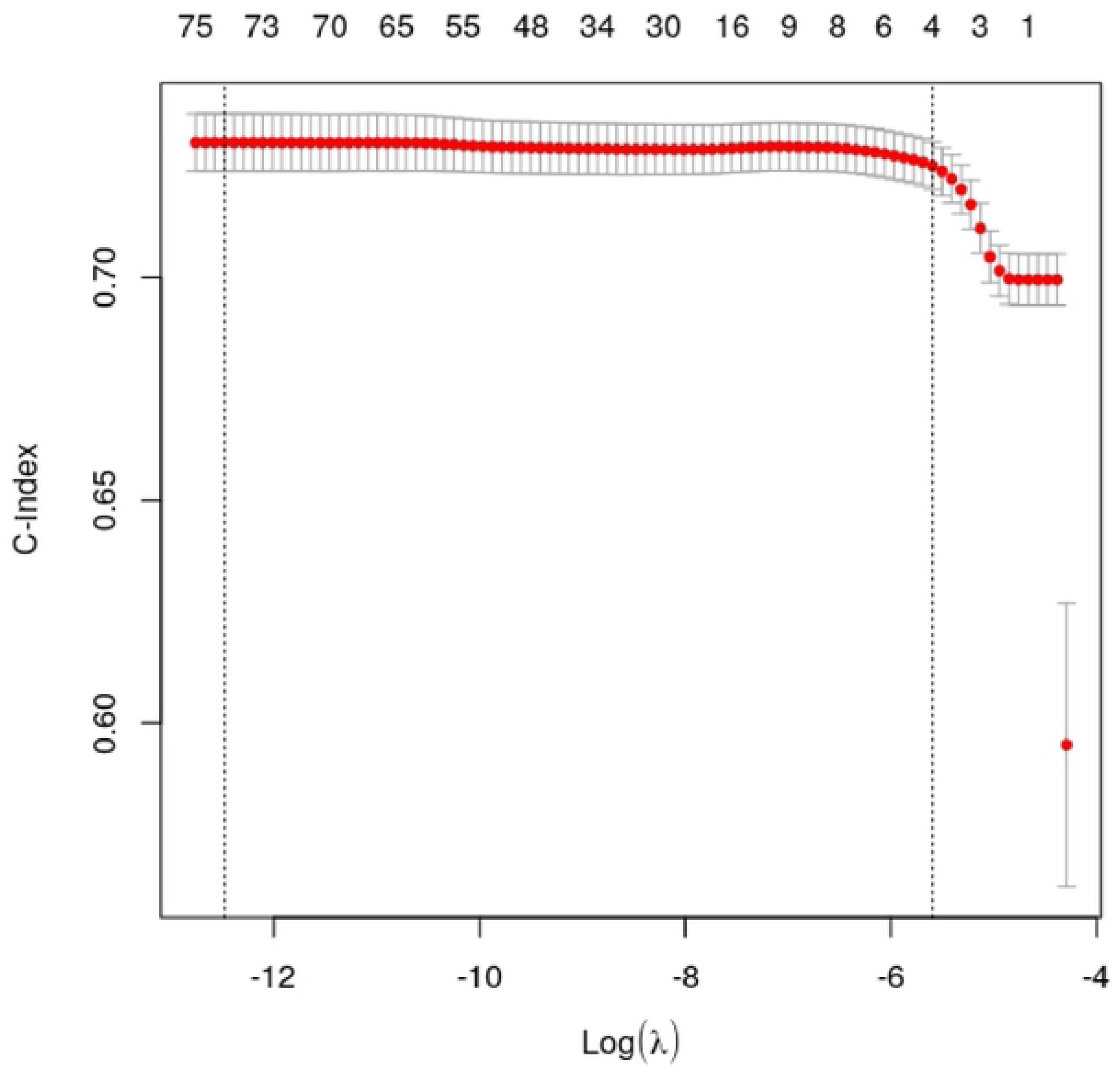
Feature Selection with diminished returns leading to Min Model.

A Lasso model average hierarchy is displayed in Supplementary Figure S1, with 16 selected features displayed in red. This model uses the optimal lambda selected from cross-validation. The tree is constructed using the hierarchical clustering algorithm to group sensor features, such that features in similar branches of the tree have similar contributions. At the center of the tree are the acceleration magnitude sensor features – enmoTrunc and enmoAbs, plus MAD and MPD. The red ones are stronger, so enmoTrunc and MPD sensor features, among the discriminating ones, are the best candidates for continuous features. These measure mean and standard deviation of average acceleration, shown to be strongly correlated with intensity of activity [22]. ENMO uses the magnitude of the acceleration and MxD uses the signal of the accelerometer.

### Min (minimum) Models

Feature selection implies a parsimonious model might be equally accurate and thus more practical, since requires less input and less compute. Hence, we explore the stepwise model strategy that utilizes small numbers of features. We denote this the *Min Model*, as shown in the flowchart in Figure 1. Such models include demographics and selected continuous features. For practical outcomes, we also include risk factors that are easy to change (especially modifiable), such as smoking and alcohol, health (general) and obesity (BMI). Min Model values are given in Table 4. We rank order the top 10 features, after considering all 76 features.

**Table 4.**
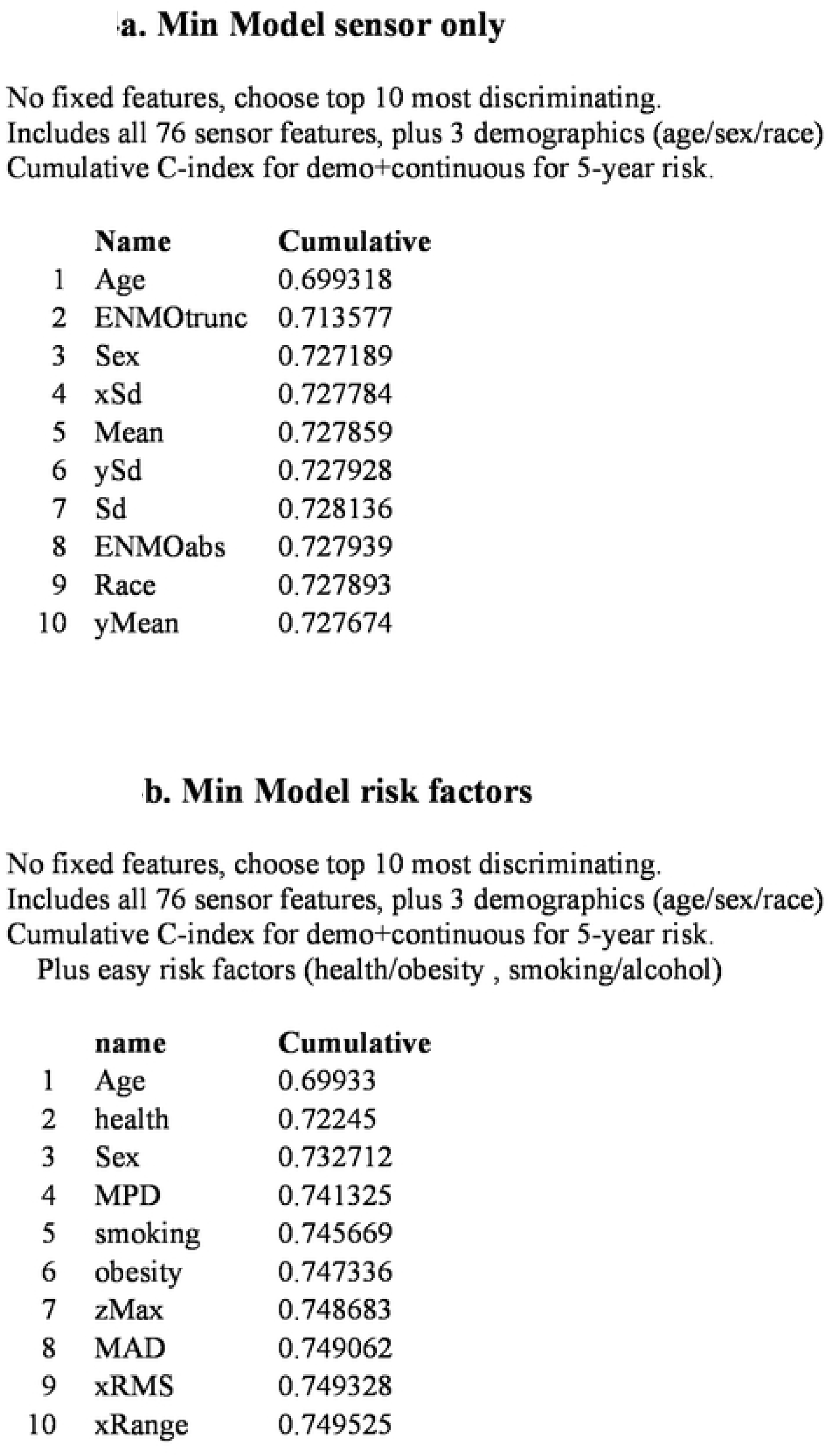
Min Model results sensor only, with Cumulative C-index rankings.

|  | **Name** | **Cumulative** |
| --- | --- | --- |
| 1 | Age | 0.699318 |
| 2 | ENMOtrunc | 0.713577 |
| 3 | Sex | 0.727189 |
| 4 | xSd | 0.727784 |
| 5 | Mean | 0.727859 |
| 6 | ySd | 0.727928 |
| 7 | Sd | 0.728136 |
| 8 | ENMOabs | 0.727939 |
| 9 | Race | 0.727893 |
| 10 | yMean | 0.727674 |

|  | **name** | **Cumulative** |
| --- | --- | --- |
| 1 | Age | 0.69933 |
| 2 | health | 0.72245 |
| 3 | Sex | 0.732712 |
| 4 | MPD | 0.741325 |
| 5 | smoking | 0.745669 |
| 6 | obesity | 0.747336 |
| 7 | zMax | 0.748683 |
| 8 | MAD | 0.749062 |
| 9 | xRMS | 0.749328 |
| 10 | xRange | 0.749525 |

If only selected continuous features are considered, then the cumulative effort on model accuracy is given in Table 4a for this minimum set. The top accelerometer feature is ENMOtrunc, acceleration magnitude correlated with activity intensity, truncated to zero for negative values [22]. With the significant demographic variables Age and Sex, the C-index is 0.727 for Min Model, rounding to 0.73, the same as the Max Model. The remaining top 10 had little extra effect, these features include other mean and standard deviation of acceleration.

When easy risk factors are also included, acceleration magnitude features improve C-index. As shown in Table 4b for Min Model, health and MPD beat smoking and obesity (BMI). This result is comparable to the UKB mortality study with categorical features only, where the most predictive features were self-reported health and walking pace [23]. The acceleration magnitude sensor features represent an objective passive measure replacing subjective walking pace. With general health included, MPD increases C-index to 0.74 using Mean Power Deviation, while slightly beating the traditional risk factors as did the self-reported walking pace [5].

### Demographic Independence

For sensor features to provide clinical utility beyond known demographics and easy risk factors, they must provide orthogonal support, for model accuracy independent of demographics. Following the original gait speed study [4], we generated the curves of 98% percentile survival time as functions of ENMOtrunc against Age and Sex. Death events are about 2% of the cohort. These mortality curves are shown in Figure 5, demonstrating sensor measures provide independent value. Higher ENMO values predict longer survival, independent of age and sex. The other acceleration magnitude features (MPD/MAD) have similar independence graphs. So the activity intensity predictor ENMO predicts mortality risk -- higher magnitude is lower risk.

**Figure 5.**
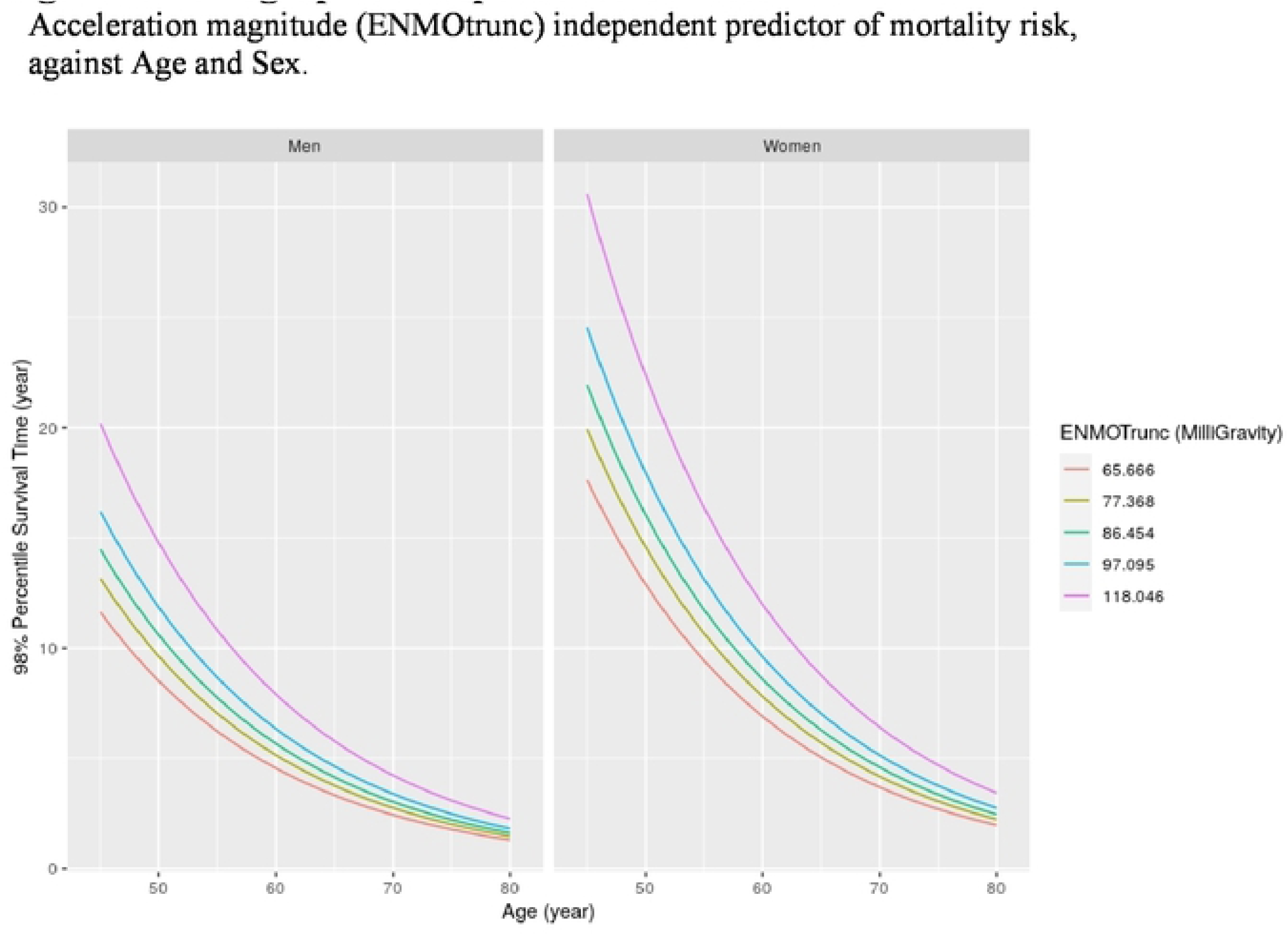
Demographic Independence Curves for sensor records.

### Geographic Variation

Over the entire cohort of 100K participants, the predictive model gives 0.73 C-index for 5-year mortality. However, it is worth noting that different populations have different accuracies using the same models. To evaluate this, we computed different statistical models (Lasso, Stepwise), then computed the C-index independently for each assessment center in 22 cities across the UK. Accuracy varies widely, as shown in Supplementary Figure S2. For example, the Lasso model averages 0.77 but ranges from 0.73 to 0.84, Lasso Continuous averages 0.73 but ranges from 0.66 to 0.77 C-index. With just Continuous, like the Min Model, Glasgow and Edinburgh have 5-year 0.77 C-index. The original mortality study without sensors had its highest reported scores at the Scottish sites as well [23]. Their analysis with categorical features only showed Glasgow and Edinburgh had 0.80 C-index with 13 questions while our categorical only is 0.81 and 0.80 with 17 questions that represent a superset of theirs.

### Intensity versus Volume

As noted, physical activity is traditionally measured by volume profile. So individuals with lower mortality have more moderate-to-vigorous activity and less sedentary activity. That is, the duration of activity is considered more important than the intensity itself. These measurement concepts for physical activity are international standards [24]. Such studies of physical activity commonly utilize wearable sensors since they are specific measures within limited periods [2]. The participants can thus be relied upon to wear the devices all day, so the studies assume 10 hours per day of wear time during normal activities. For effective usage in daily living, the patients must continuously wear a medical quality sensor device. In contrast, our methods assume a single 6MWT per day, so 6 minutes rather than 600 minutes, two orders of magnitude less sensor data. Our methods enable the usage of cheap smart phones, since often carried while walking yet having adequate accelerometers for predictive models of pulmonary function [16].

With cardiopulmonary patients, intensity is more important than duration, as shown in large meta-analysis studies [25]. Our model prediction relies upon walking intensity in short bursts being an effective surrogate for activity intensity over whole days. Intuitively, walking is the unique physical activity, which ranges in intensity from vigorous (fast) to sedentary (slow). Brisk walks are nearly as vigorous as running and shuffling walks are nearly as sedentary as standing.

Another confirmation of walking intensity as an effective surrogate for activity volume is shown by our Lasso model average hierarchy in Supplementary Figure S1, with the most discriminating selected sensor features displayed in red. The center of the tree is the acceleration magnitude features such as ENMO and MPD. In a single central branch for selected features are MCR, Mean Crossing Rate, along with MMCR (Maximum and Minimum average Crossing Rate), the closest equivalent with walking intensity to RA (Relative Amplitude) which measures activity volume, as described below. In addition to overall average for MMCR, nearby branches include in red, the specific yMMCR and zMMCR due to walking motions. But the other 4 features in red near the center are the equivalents of RA for short bursts, which we computed for comparative purposes as special additions to biobank features. These concern the crossing rate, how often the acceleration changes from above the mean to below the mean and vice versa, which we previously utilized for predictive models of fall risk in national cohorts [10].

### Comparing Accuracy to Concurrent Study

The UK Biobank accelerometer dataset has also been analyzed by a concurrent study which used cooked data to analyze the activity volume [26], rather than using the raw data as we did to analyze the walking intensity. This study took the 5 second averages from Biobank field 90004 and further averaged over 1 minute intervals. Our study used raw signals from field 90001, which can detect characteristic motions of walking intensity, since 1 minute contains 6000 data points rather than only 1. Our Min Model achieves the same C-index accuracy of 0.72 as their study for continuous features with demographics, although their analogue of ENMOtrunc called Relative Amplitude requires 100 times more minutes per day. (RA compares the highest 10 hours of activity to the lowest 5 hours, so need 600 minutes rather than 6 per day). Since they are measuring volume (quantity) of physical activity, they average the sensor records to 1 value per minute of accelerometer data. Since we are measuring intensity (quality) of physical activity, we use the raw data at 100 Hz which is 6000 values per minute. So each day, we measure 100 times less minutes with 10 times more samples. Due to their requirement of at least 3 days with at least 10 hours per day sensor records, they excluded 21K participants while we excluded 3K participants, so we have 7 times more inclusions for measured participants.

Given the geographic variation, we could focus solely upon the highest accuracy at the Scottish sites of Glasgow and Edinburgh, as did the original UK Biobank mortality study using self-reported questionnaires only. There we achieve 5-year 0.77 C-index with the Min Model. This is 5 points higher than the concurrent study using activity volume, despite using 30 minutes instead of 30 hours of sensor input, per participant per week of sensor measure. In considering the Max Model with categorical as well as continuous features, the concurrent study listed similar categorical features to ours but did not explain the derivation. We give the Biobank fields used to extract our categorical features in Supplementary Table S1, with summary statistics for participants and values.

In our study, we used Date fields giving physician consensus for disease diagnosis, more detailed from more sources than the participant self reported answers used in the concurrent study. This enables a fair comparison between categorical and continuous features, both up to date at the time of sensor records. The self reported disease features are from participant registration, which is 6 years before the sensor records (2008 versus 2014). So their sensor features are more recent than their categorical features, and hence more accurate for mortality prediction than is actually correct, similar to 5-year versus 10-year mortality risk. Their conclusion that objective measures with sensor features improve prediction performance of risk factors thus may be flawed, a confounded artifact of when data was gathered.

## DISCUSSION

Measuring activity via walking intensity has become standard practice for clinical visits, where gait speed measures short walks. Detailed meta-analysis showed gait speed independent of age/sex [4], with pooled C-index close to 0.72 model accuracy for 5-year risk. Objective physical activity (OPA) measures the “quantity” of physical activity, such as total amount of moderate-to-vigorous physical activity. This requires sensor devices be worn all day, so a concurrent study [26] of the UK Biobank sensor dataset showed the highest predictor was relative amplitude (RA), ratio of most active 10 hours of average acceleration to least active 5 hours. The C-index was 0.72, with RA plus age/sex for 5-year risk, based upon 600 minutes per day of sensor records.

A walk test measures “quality” (intensity) rather than “quantity” (volume). Our previous work showed accelerometer sensors in carried smartphones can digitally model physical distance [27] and oxygen saturation [28] from Six Minute Walk Test (6MWT). We also showed the pulmonary models similarly worked with carried smartphones during daily living [16]. The logistic advantage of 6 minutes walking intensity is two orders of magnitude less frequent sensor input, using ENMO for quality versus RA for quantity. This makes it possible to effectively utilize smart phones instead of wearable sensors for predictive models.

The Min Model with fewer sensor features holds at the same 0.72 for 5-year risk. Using continuous features only without categorical features, our Max Model yields 0.73 C-index for 5-year risk, with greater accuracy in earlier years yielding 0.76 for 1-year risk. We note model accuracy varies by local sites, as shown in Supplementary Figure S2, with 0.77 for 5-year at Glasgow and Edinburgh, where the original mortality study using self reports also did best [23]. Our digital measures of walking intensity are better in quality for predictive power versus traditional modifiable risk factors such as smoking and obesity. This objective result confirms the previous subjective UK Biobank study using self reports of walking pace [5].

We are involved in planning the physical activity study for the US Precision Medicine Initiative (All of Us Research Program). This cohort is projected to become the largest national cohort with more than 1M participants, close to half already registered. These participants are being recruited to be representative of the national population, which is far more diverse in the US than in the UK. All agreeing participants would be longitudinally measured on their personal smartphones, both larger and longer than our mortality study as well as directly utilizing phone sensors for the measurement study.

Our previous work showed accelerometer motion sensors in cheap smart phones can capture equivalent model input for gait analysis as expensive medical devices. This is particularly important for health equity purposes, given populations at highest health risk are often the most under-resourced—persons most likely to have cheap phones rather than wearable devices would benefit most from easy assessment. Phone apps could record six minutes of consecutive walking during daily living, then compute predictive models for risk stratification via population analysis [11]. Major cohort studies using self reported status have shown that cardiovascular health is strongly correlated with physical activity, largely independent of the socioeconomic level of the country of the participants [29]. Thus our results from high-income countries may well be directly applicable to low-income countries as well. Healthy longevity could be facilitated for all adults possessing cheap phones, using the minimum model to assess gait status, computed on their phones for the maximum privacy. Implementing effective healthcare infrastructure requires continued research into screening populations with ubiquitous sensors [30].

## MATERIALS AND METHODS

### Ethics Statement

This study analyzes datasets provided by UK Biobank, with subjects identified only by participant number. This Biobank is a national resource in the United Kingdom, providing datasets to international researchers who have approved projects. Our project entitled “Predictive Models of Mortality Risk from Passive Monitors measuring Physical Activity” is approved with ID 45178. This enabled us to download datasets with selected portions of their complete database, each dataset was approved by the Biobank as was each investigator including all of the authors. UK Biobank supports extensive human subjects protection including written informed consent from each participant. The signed Materials Transfer Agreement between University of Illinois and UK Biobank specifies that we will abide by all their ethical standards.

### Study Participants

UK Biobank is a prospective study with over 500,000 participants aged 40-69 years [8]. These participants were recruited during 2006-2010 from 22 assessment centers throughout the UK. The study is longitudinally collecting participants’ information, including data from questionnaires (self reports), physical measures (laboratory tests), and accelerometers (sensor records). It is representative of the national population for demographic and geographic considerations, although the entire cohort shows less disease and more education than the UK population at large [17]. Within the entire cohort, traditional risk factor associations agree for mortality outcomes with nationally representative cohort studies [31]. Thus the cohort dataset for sensor analysis is uniquely suitable for predictive models. UK Biobank provides accurate datasets for sensor input with physical activity and status output with health outcomes [19].

Our study focuses on the subset of 103,683 participants who agreed to wear a wrist-worn triaxial accelerometer, an Axivity AX3 sampling at 100Hz, continuously for 1 week [9]. These participants were aged 45-79 when data was collected in 2013-2015. We implemented inclusion/exclusion criteria shown in Figure 2. We exclude 257 participants for insufficient device wear time. Our analysis focused on walking intensity, so participants must have sufficient length of steady walking, as defined below. The Biobank software [19] divides sensor data into non-overlapping 30-second windows with activity labels. The Biobank software divides sensor data into non-overlapping 30-second windows with activity labels. These are highly accurate, due to careful derivation from training set of representative participants who wore head-mounted cameras to visually identify activities. We included any participant with at least one session of steady walking, defined by 12 consecutive walking windows. Only windows labelled as walking were considered input data for feature extraction. We exclude 2758 participants for insufficient walking, which with other minor exclusions, yields total 100,655 participants for our analysis. There were 2048 included deaths from UK Biobank field 40023, derived from the National Death Registry, which is a comprehensive curated dataset. We analyzed all-cause 5-year mortality, with sensor records from Jun 2013 to Dec 2015 and deaths until Dec 2019 to avoid COVID-19. For highest accuracy, our analysis used all qualifying participants, after trying different subsets including different age ranges.

### Steady Walking in Daily Living

Walk tests are widely used to clinically evaluate status of cardiopulmonary patients. A standard assessment is the Six Minute Walk Test (6MWT), where a patient walks back and forth in a corridor for six minutes and their walked distance indicates their health status [6]. With COPD patients, this period is long enough so patients slow down in correlation with their status determined by spirometry [32]. Such walk tests are also used for CHF patients [33], who also exhibit Shortness of Breath on Exertion (SOBOE) [34]. We have previously shown with such cardiopulmonary patients that accelerometer sensors can measure slowdown/speedup with clinical accuracy, for predictive models of 6MWT distance and pulmonary function [27, 16]. These were clinical experiments with COPD/CHF patients who performed 6MWT in hospital rehabilitation, with carried smartphones recording accelerometer sensors.

There is no current standard for walk tests during daily living. We chose 6 minutes as empirical lower bound for cardiopulmonary slowdown during *steady walking*, with relaxed criteria to allow longer periods. During daily living, a person may walk more slowly than when they are pushing hard during walk test, so it might take longer for them to experience SOBOE. Thus we require 12 consecutive walking windows to be the “equivalent of 6MWT”, and include all such labelled windows for included participants with at least 1 such session. For example, 4 consecutive walking windows would be excluded, while 12 consecutive windows or even 20 would be included. All participants had 1 session of 6 minutes continuous walking during the 1 week, although only 10% walk half an hour in 6 minute sessions as shown in Figure 6.

**Figure 6.**
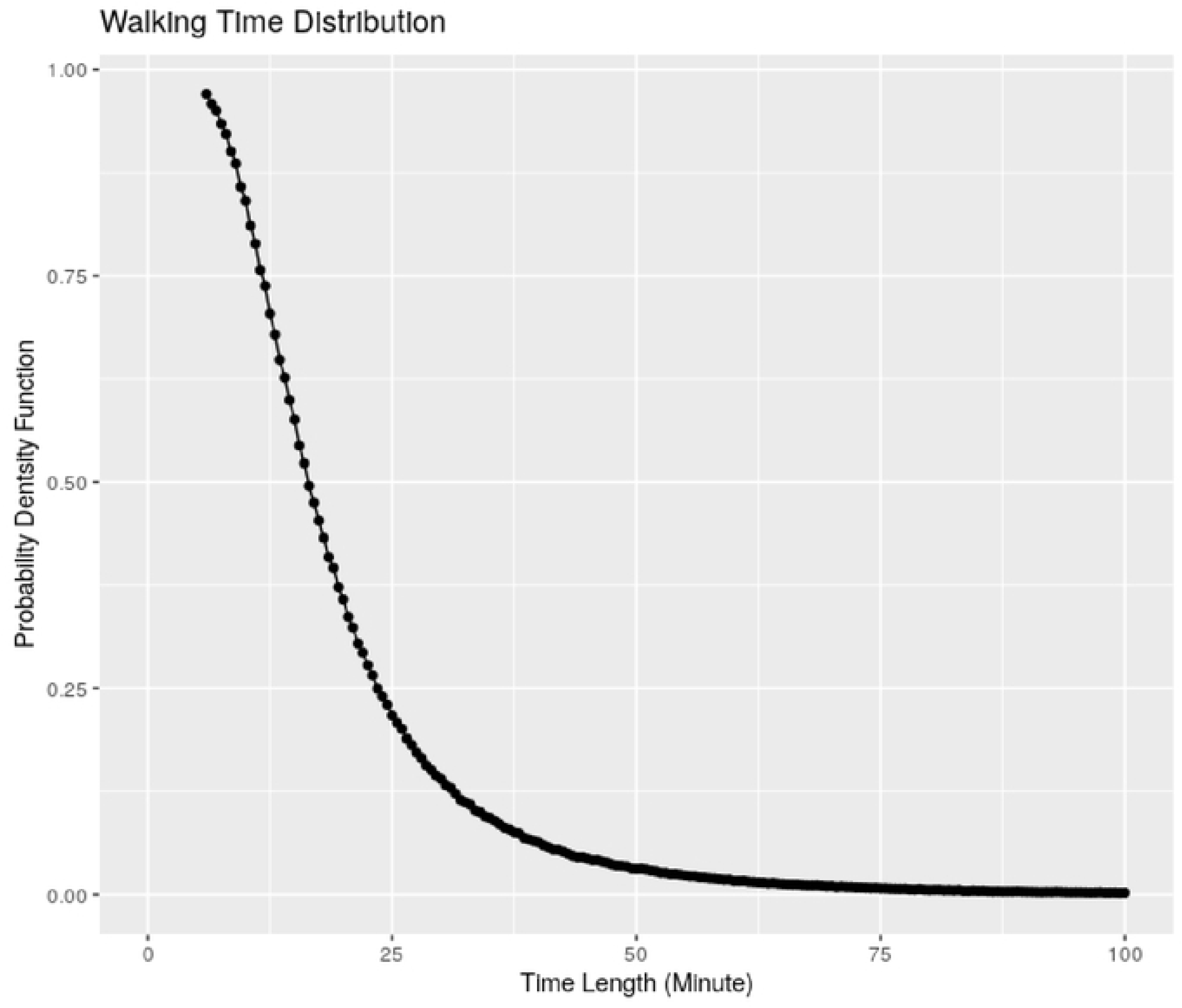
Walking Distribution of Labelled Sessions during Daily Living.

### Mortality Predictors: Traditional Categorical and Accelerometer Continuous

UK Biobank collected questionnaires during 2006-2010, when participants registered. We extract categorical features based upon self-reported answers and laboratory tests. We select 20 questions to characterize health status, including 7 Advanced Disease Conditions, 10 Modifiable Risk Factors, 3 Demographics. These are grouped into categories in Table 1. The original analysis for mortality risk using only categorical features found 13 features to be most discriminating [23], our list includes theirs. Supplementary Table 1 gives summary statistics.

Accelerometer data is collected by Axivity AX3 wrist-worn accelerometer, which collects 100Hz triaxial signals [9]. We follow the provided Biobank methods to extract features from raw data [19]. Our previous work showed raw sensor data was needed to predict status of pulmonary function in clinical studies with cardiopulmonary patients [16]. Model input was 6MWT sensor records, so more data points are needed than simply average sensor data over a labelled walking window.

Every 30 second time window is an epoch for measuring physical activity with single label, yielding 3000 3D acceleration points. The total number of possible epochs over 7 days is 20160. In each epoch, we extract a 76-dimensional feature vector, where each feature describes certain characteristics of motion patterns. Signal features are derived from time domain and frequency domain [35, 36]. We use only features from time domain, since measuring walking intensity over time periods. We used 38 features from the Biobank software time domain [19], and computed another 38 features from their frequency domain following our previous work [16]. The 76 sensor features are listed in Table 2, while Supplementary Table 2 gives their formulas.

We added dimensional data, such as x-y-z, plus computing our own features from the frequency domain of Biobank software^13^. Our new features include those useful in other studies. These included RMS (root mean square) from our prior work [16], which is computable from FFT. Total activity count (TAC) is the overall feature provided by commercial fitness devices such as research standard Actigraph GTX-3. We computed comparisons of most active periods to least active, such as MMCR (Maximum and Minimum average Cross Rate) and MCR (Mean Cross Rate), similar to physical activity profiles from activity volume methods. Full formulas for sensor features are given in Supplementary Table 2. The major features are defined in terms of ENMO, mean acceleration correlated with activity intensity such as walking versus standing. These include MPD and MAD, Mean Power Deviation and Mean Amplitude Deviation of accelerometer magnitude signals [37].

### Sensor Data Processing

The raw data was collected into 30-second windows over the entire week of recording, each window contained 3000 3-axis motion samples from field 90001. This comprised 25 terabytes. We analyzed this dataset using the Biocluster2 at the Carl R. Woese Institute for Genomic Biology, which has 72 nodes of Xeon Gold 6150s with 2 cores each of 2.7 GHz and 4GB memory. We computed 1037 batches of instances, where each batch consists of 100 instances. These instances covered the participant sensor record for each of almost 103,700 participants.

The total processing time for feature extraction was 3100 hours of compute time. It takes about 3 hours for each node to process each batch. The typical sensor processing used 50 nodes on the shared cluster, so the total real time was about 62 hours, or about 3 days. The steady state of extracted features is 1.2 TB, which we kept on the cluster storage as input to run the models.

### Mortality Prediction Models and Survival Function Estimation

The prediction response is the time interval between end time of participant mortality and device wearing for sensor record. Since the outcome is time-to-event and subject to censoring, we utilize survival analysis [38]. Hence we consider the Cox proportional hazard model [39] and its penalized version [40]. We use the elastic net penalty [41], which consists of both l1 and l2 penalties of the coefficients, controlled by two tuning parameters α and λ. The method is implemented using the R package glmnet [42]. To select the optimal tuning, we consider α = 0, 0.5, 1 and use a grid of λvalues automatically selected by the glmnet package.

We then perform 10-fold cross-validation [43]. Cross-validation procedures are more stable than pre-fixing testing data since they enable all observed data to be used in evaluation steps. These are commonly used in data analytic procedures with machine learning methods [44]. The procedure 10-fold is the averaged result of performing 10 such pre-fixing procedures. In contrast, results derived from pre-fixing the testing data only once can be greatly effected by the randomness involved in choosing the test set.

We use a stratified 10-fold cross-validation approach, since the proportion of death is small (about 2% of participants). For each model with maximum follow-up length (1/2/3/4/5 years), we consider the data is randomly split into 10 equal-sized subsets. Each subset contains 1/10 of the live data (participants who are still alive or censored by the maximum follow-up time) and 1/10 of the dead data (participants who have died by the maximum follow-up time). With these 10 equal-size datasets, a single subset is utilized for testing the model and the remaining 9 subsets are used as training data. The cross-validation process is repeated 10 times with every subset used exactly once as the testing data. Finally, the 10 results from the folds can be averaged to produce a single estimation for a specific model with exact $alpha$ and $lambda$. For each $alpha$, the $lambda$ with best performance is selected from grid as model parameter. In addition to the regularized Cox proportional hazards model, we fit other models to compare their performance. We adopt stepwise selection to choose variables. With fixed variables as input, we set the prediction performance as inclusion criteria to do stepwise forwards selecting over these variables. In every step, the variable that increases the C-index the most based on the previous selected variables is included in the group. The selection runs until the increment is less than a specific threshold. We have tested over traditional predictors and accelerometer derived predictors with threshold 0.01 and 0.001, to enable model evaluation.

The Concordance Index (C-index) is used to evaluate the model performance [45]. The C-index can be interpreted as the fraction of all pairs of subjects whose predicted survival times are correctly ordered as the observed survivals, while correcting for censoring. Hence, it is more sensible than other common criteria such as the overall accuracy or the Area under the Curve.

## Data Availability

The datasets analyzed are owned and controlled by UK Biobank, a national research resource in the United Kingdom. As a public resource, all this data and more is available to interested researchers worldwide. However, all such requests must be formally approved by the Biobank and the datasets are not available otherwise.

## ACKNOWLEDGEMENTS

Qian Cheng at Salesforce AI Research advised us on predictive models of wearable sensors.

---

**Figure S1.**
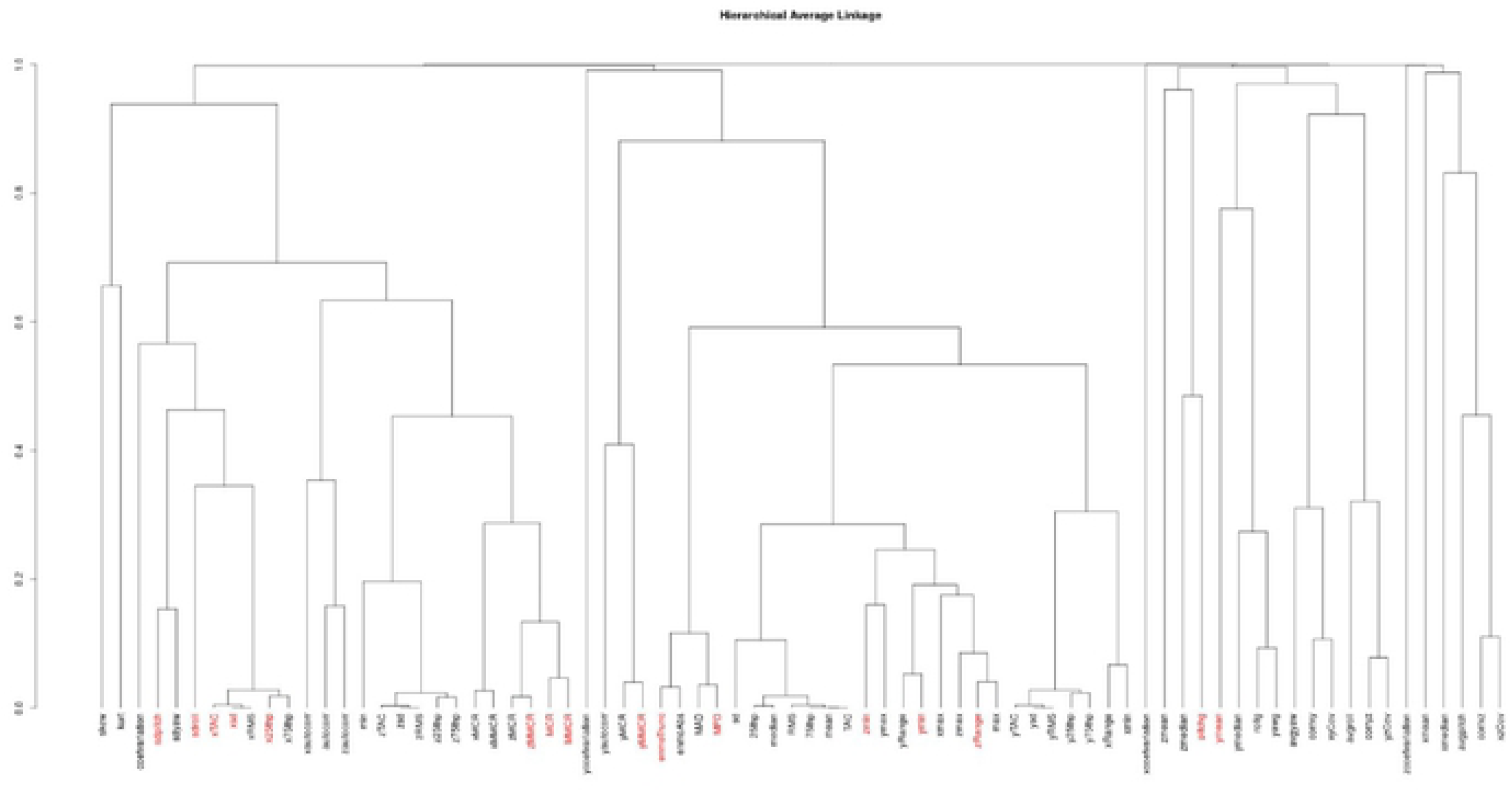
Lasso Model. Hierarchy tree average with red selected features.

**Figure S2.**
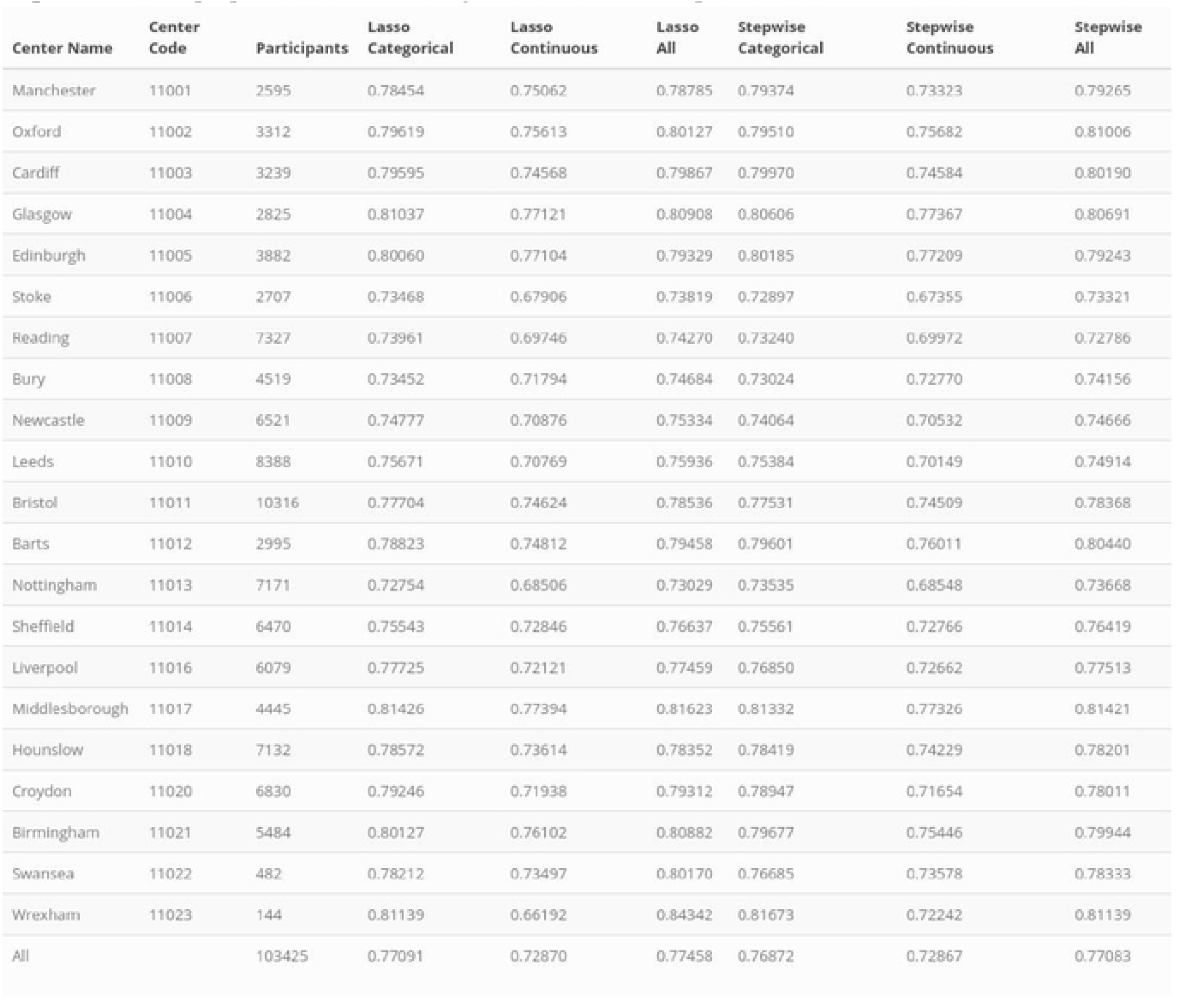
Geographic Variation of Models across Cohort sites.

**Table S1.**
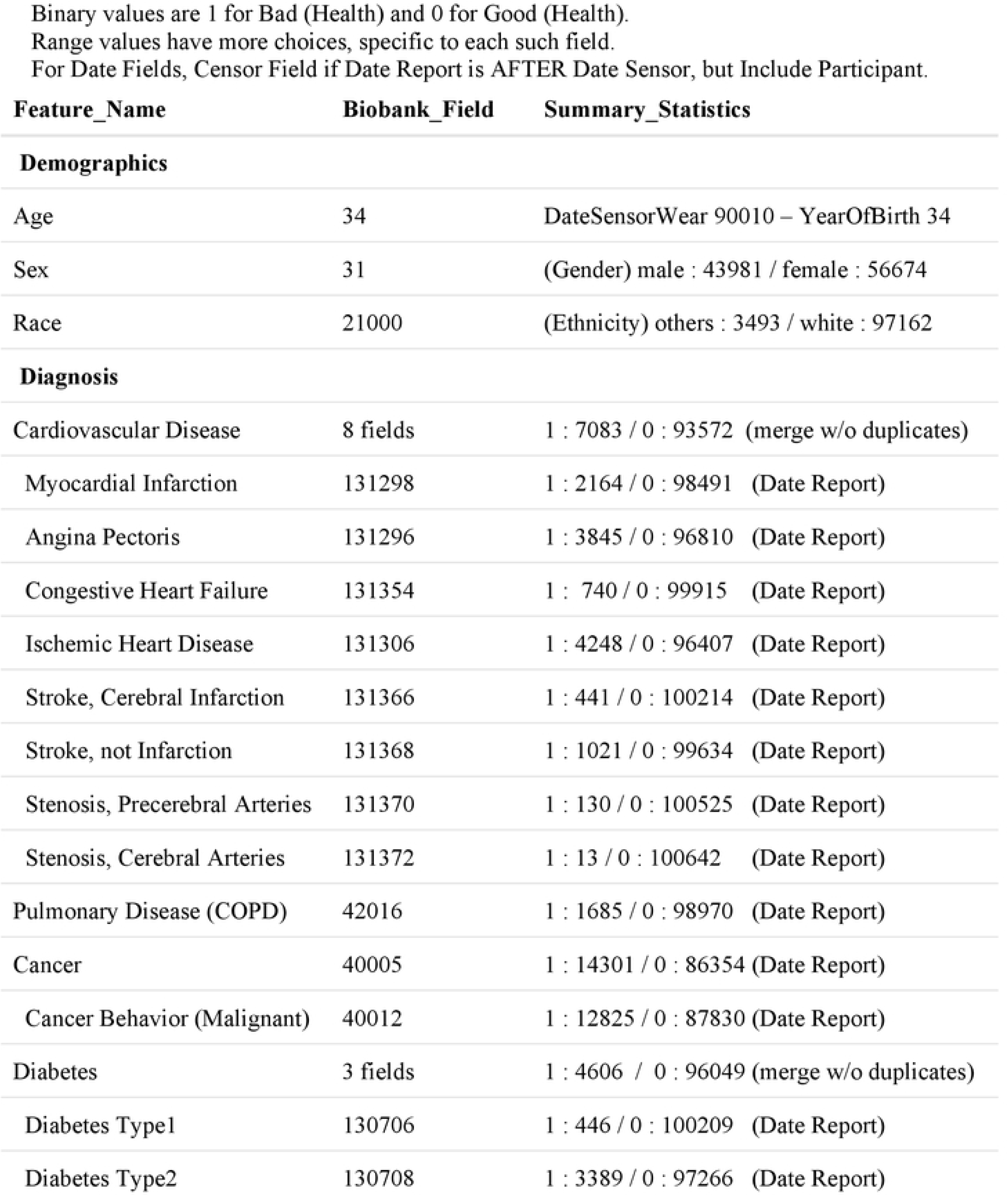
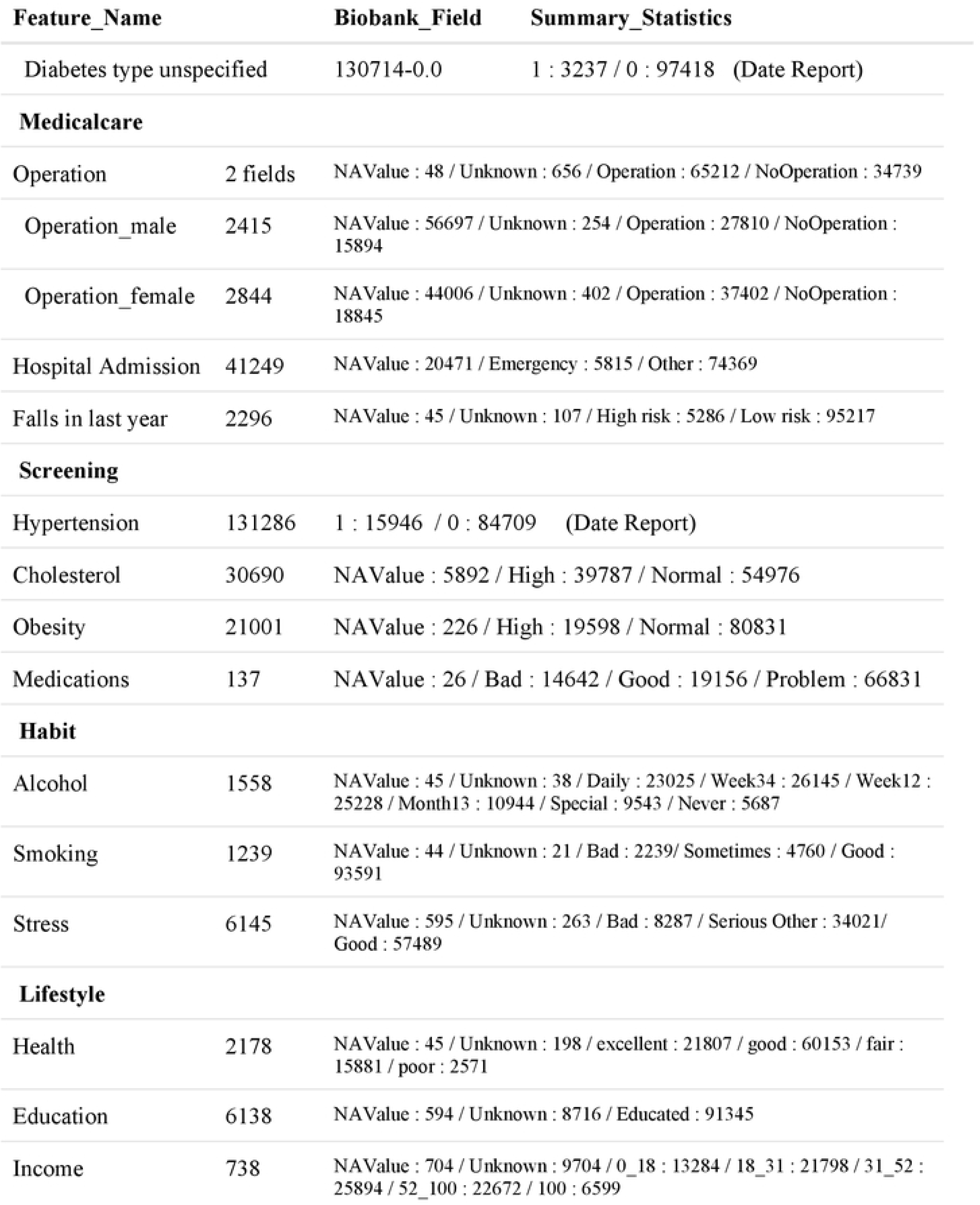
Categorical Encoding. Features from cohort dataset of 100,655 participants.

**Table S2.**
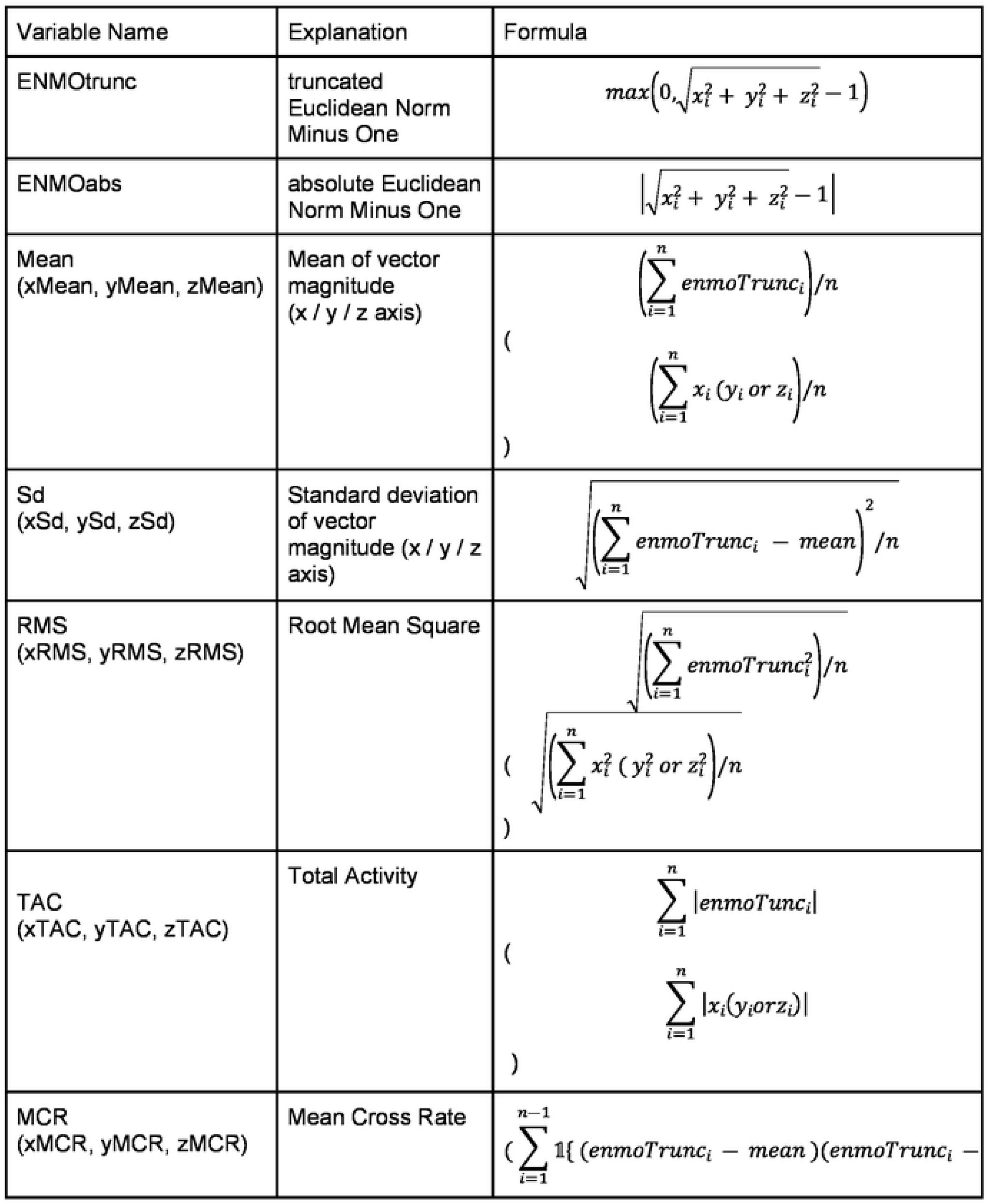
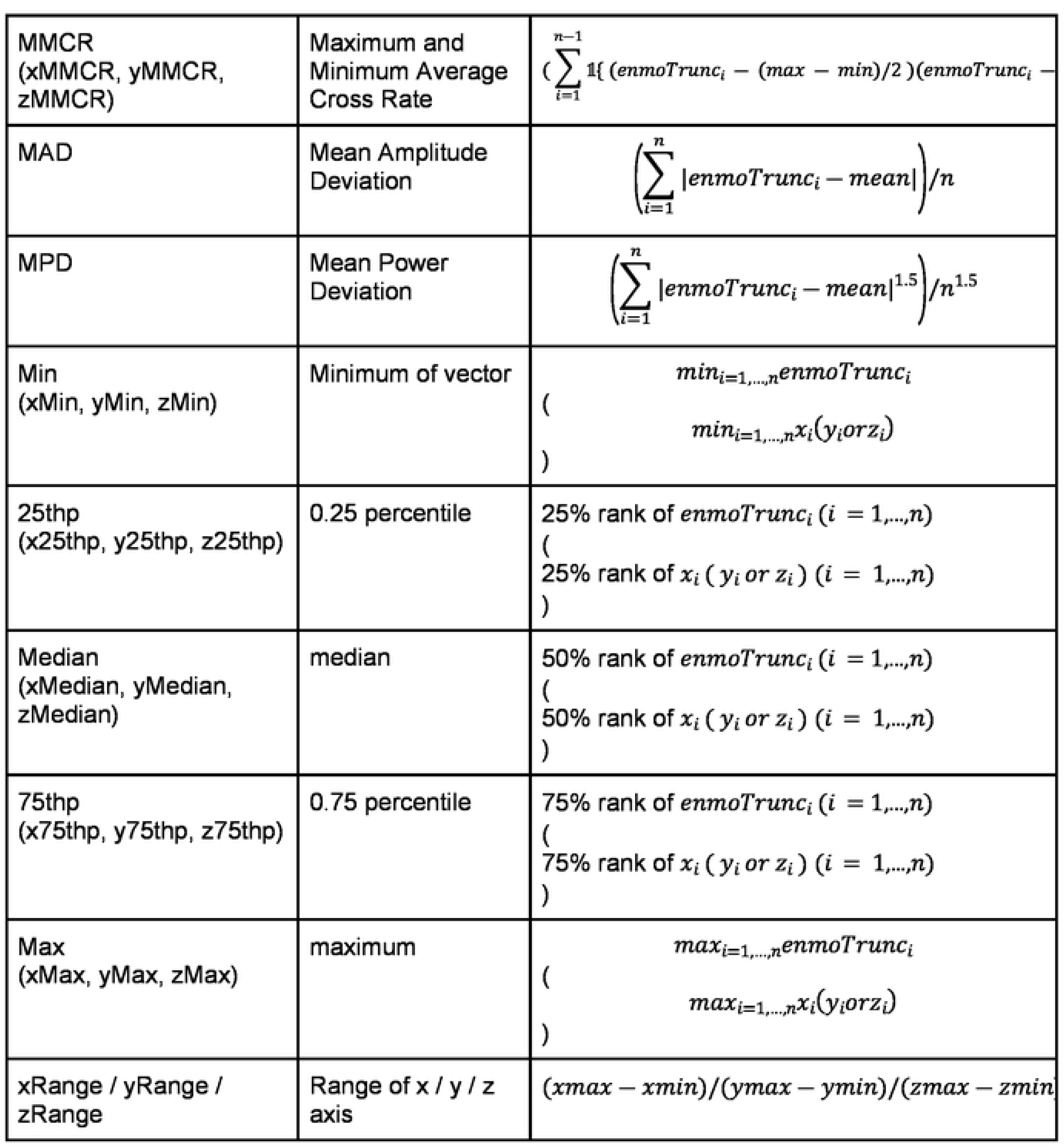
Continuous Encoding. Formulas for Accelerometer Derived Sensor Features.

**Table S3.** Marginal Performance of Top 30 of All Features ranked by C-index.

| **Name** | **C-index** |
| --- | --- |
| Age | 0.699329 |
| enmoTrunc | 0.639477 |
| MPD | 0.637168 |
| MAD | 0.630199 |
| enmoAbs | 0.629914 |
| Sd | 0.626054 |
| RMS | 0.615782 |
| 75thp | 0.612617 |
| yRange | 0.612016 |
| Mean | 0.61102 |
| TAC | 0.610951 |
| xRange | 0.61039 |
| Ymin | 0.609938 |
| Income | 0.608182 |
| Median | 0.60627 |
| Max | 0.604575 |
| Ymax | 0.603579 |
| zRange | 0.603381 |
| Xmax | 0.602969 |
| Xmin | 0.600941 |
| Zmin | 0.600338 |
| medication | 0.598063 |
| 25thp | 0.597637 |
| xRMS | 0.597255 |
| Xsd | 0.59719 |
| Zmax | 0.5958 |
| xTAC | 0.590543 |
| hypertension | 0.58908 |
| Health | 0.586785 |
| Sex | 0.584188 |
| x25thp | 0.583206 |
| x75thp | 0.581649 |
| hospital_admission | 0.580935 |
| Cancer | 0.580095 |
| Ysd | 0.579393 |
| yRMS | 0.579341 |
| Zsd | 0.577168 |
| zRMS | 0.577154 |
| yTAC | 0.572009 |
| zTAC | 0.56789 |
| heartdisease | 0.565598 |
| y75thp | 0.565253 |
| y25thp | 0.562617 |
| stress | 0.559431 |
| z75thp | 0.556267 |
| z25thp | 0.555108 |
| obesity | 0.547041 |
| min | 0.546833 |
| alcohol | 0.545644 |
| education | 0.542326 |
| diabetes | 0.536643 |
| sdpitch | 0.533721 |
| sdyaw | 0.531501 |
| smoking | 0.530064 |
| pitchg | 0.529827 |
| operation | 0.526503 |
| LungDisease_COPD | 0.526217 |
| skew | 0.522987 |
| cholesterol | 0.52042 |
| xautocorr | 0.516339 |
| falls | 0.513014 |
| sdroll | 0.512742 |
| ymedian | 0.509274 |
| xyCov | 0.508795 |
| corrxy | 0.507807 |
| ymean | 0.507165 |
| avgyaw | 0.50679 |
| yzCov | 0.506411 |
| corryz | 0.505973 |
| xmean | 0.505578 |
| zmean | 0.505249 |
| avgpitch | 0.504397 |
| kurt | 0.50365 |
| yautocorr | 0.503615 |
| coefvariation | 0.503595 |
| xMCR | 0.503576 |
| Race | 0.50246 |
| zmedian | 0.502363 |
| MMCR | 0.498242 |
| zcoefvariation | 0.49763 |
| rollg | 0.496389 |
| xMMCR | 0.495558 |
| corrxz | 0.494586 |
| ycoefvariation | 0.494541 |
| xzCov | 0.492141 |
| avgroll | 0.491459 |
| autocorr | 0.49085 |
| xcoefvariation | 0.490604 |
| yawg | 0.486496 |
| yMCR | 0.484588 |
| xmedian | 0.484326 |
| zautocorr | 0.482311 |
| yMMCR | 0.478037 |
| zMMCR | 0.476288 |
| MCR | 0.47558 |
| zMCR | 0.469512 |

